# Incidence and Predictors of Burnout in Healthcare Postgraduate Trainees Under a Widespread, High-Demand Sanitary Crisis: A Longitudinal, Observational Study

**DOI:** 10.64898/2026.05.07.26352624

**Authors:** Thais Ferreira Costa, Rebeca da Nóbrega Lucena Pinho, Nayane Miranda Silva, Adriana Ferreira Barros Areal, André de Matos Salles, Andrea Pedrosa Ribeiro Alves Oliveira, Carlos Henrique Reis Esselin Rassi, Ciro Martins Gomes, Dayde Lane Mendonça da Silva, Fernando Araújo Rodrigues de Oliveira, Isadora Jochims, Ivan Henrique Ranulfo Vaz Filho, Lucas Alves de Brito Oliveira, Marta Alves Rosal, Mayra Veloso Ayrimoraes Soares, Patrícia Shu Kurisky, Viviane Cristina Uliana Peterle, Ana Paula Monteiro Gomides, Cezar Kozak Simaan, Veronica Moreira Amado, Cleandro Pires de Albuquerque, Licia Maria Henrique da Mota

## Abstract

**Background:** High-demand sanitary crises, such as the COVID-19 pandemic, impose a high burden on healthcare professionals, increasing their risk of burnout. Healthcare postgraduate (HCP) trainees compound the general healthcare professional workforce and may face unique risks and challenges. This study aimed to evaluate the incidence of burnout and identify its predictors among healthcare postgraduate trainees during a high-demand sanitary crisis.

**Methods:** A longitudinal observational study was conducted during the pandemic among healthcare postgraduate trainees from 67 Brazilian healthcare institutions. Participants were assessed at baseline (July to September 2020) and after an 18-months follow-up. Individuals with burnout at baseline were excluded. Several questionnaires, including the Oldenburg Burnout Inventory (OLBI) and the depressive disorder PHQ-9 scale were applied. Associations between baseline characteristics and the development of burnout were analyzed using chi-squared and t tests, and log-binomial regression. The study received ethical approval (CAAE: 33493920.0.0000.5558).

**Results:** A total of 313 participants were included; mean (SD) age: 28.2 (4.6) years; 80.1% (n=250) were biological females; 58.5% (n=183) whites; 51.1% (n=160) physicians; 12.5% (n=39) nurses; 36.4% (n=114) other HCP trainees; 47.9% (n=150) had depressed symptoms at baseline. Burnout incidence rate [95% CI] was 202.9 [166.5, 239.3] cases per 1000 person-years. In bivariate analyses, depressive mood at baseline predicted future burnout (relative risk [95% CI] = 2.14 [1.49, 3.08]; p<0.001), while older age (mean difference, MD [95% CI] = 1.10 [0.16, 2.09] years; p=0.029), higher autonomy (MD [95% CI] = 0.57 [0.10, 1.04] on a 10-point visual numerical scale, VNS; p=0.018) and adequate professional training (MD [95% CI] =0.85 [0.30, 1.40] on VNS; p=0.003) showed protective effects. Sex, race and weekly workload could not predict burnout. In multivariate analyses, depressive symptoms at baseline remained independently associated with higher risk of burnout (risk ratio, RR [95% CI] = 1.84 [1.26, 2.71]; p=0.002), while having adequate professional training showed a protective effect (RR [95% CI] = 0.61 [0.43, 0.87]; p=0.007).

**Conclusions:** Very high incidence of burnout among HCP trainees was observed under a global sanitary crisis. Depressed mood at baseline was the most relevant predictor of subsequent burnout. Providing mental health support for HCP trainees in future widespread sanitary crises seems advisable to preserve the workforce.

## Introduction

Burnout syndrome describes a state of physical and mental exhaustion related to caregiving activities and can affect job performance as well as worker well-being(1). It is considered a major public health issue leading to significant physical and emotional impacts (2). According to Maslach, burnout comprises three key components: emotional exhaustion, depersonalization, and reduced personal accomplishment (3). Emotional exhaustion describes a state of being mentally and physically drained, due to prolonged stress or overwork. Depersonalization is marked by a detached, negative attitude, where individuals may distance themselves emotionally from their work or from others. Finally, reduced personal accomplishment reflects a diminished sense of efficacy, where individuals perceive their work as ineffective and unfulfilling (3).

Burnout imposes a significant occupational hazard to professions that demand prolonged and intense interpersonal engagement, such as healthcare, education, and other service-oriented fields. These roles often require professionals to exhibit exceptional dedication, generosity, and self-sacrifice toward their clients, patients, or students, increasing their susceptibility to burnout (4).

In 2020, a global sanitary crisis brought about by the COVID-19 pandemic significantly disrupted healthcare systems and workplace organizations worldwide, presenting unprecedented challenges for institutions and individuals (5). In Brazil, the first verified case of Coronavirus Disease 2019 (COVID-19) was reported in São Paulo, on February 26, 2020 (6). Since then, according to official data from the Ministry of Health, until September 2025, the country had recorded more than 39 million accumulated cases and more than 716 thousand deaths (7).

In the context of this widespread sanitary crisis, healthcare professionals under training, encompassing medical and other health specialties, residents and fellows, who were assigned to treat patients with the disease, experienced intense physical and psychological pressure (8). Residency programs are structured as in-service training under supervision, aimed at developing professional skills and competencies (9). A systematic monitoring on the quality of life of these professionals is needed given their extensive and intense workload (60 h per week in Brazil) (10). Factors such as staff shortages, prolonged use of personal protective equipment, and sleep deprivation (even before the sanitary crisis) predisposed this group to the development of mental health disorders (8), including burnout syndrome. Furthermore, a longitudinal investigation among internal medicine residents revealed a direct correlation between elevated levels of burnout and an increased propensity for reporting medical errors (11)

Burnout and its impact among physicians and other healthcare trainees in learning environments remains insufficiently studied. The existing literature on this subject is limited, despite the crucial need to understand the effects of high-demand sanitary crises on these healthcare professionals. This understanding is essential to develop strategies that can help address the problem.

The aim of the present study was to assess the incidence and predictors of burnout among healthcare postgraduate trainees during a high-demand sanitary crisis.

## Materials and methods

This was a longitudinal, prospective, observational cohort study conducted under the high-demanding sanitary milieu presented by the COVID-19 pandemic. The study protocol as well as the findings of a closely related, cross-sectional study within this same research line have already been published (12) (13).

Eligible participants were adults (aged 18 years or older) under professional training within medical and other health care professions residency and fellowship programs, directly involved in patient care during the pandemic sanitary crisis. The only exclusion criterion was already presenting burnout at baseline.

Recruitment was carried out through various channels, including email, social media messages, posters displayed in hospitals, and university hospitals intranets. These materials featured QR codes linking directly to the study forms. Initial data collection occurred between July and September 2020.

Links to questionnaires were first sent to 7,215 residents from 40 university hospitals managed by EBSERH (Empresa Brasileira de Serviços Hospitalares), a public company under the Ministry of Education responsible for overseeing federal university hospitals. Participation, however, was also open to healthcare postgraduate trainees from other training institutions across Brazil.

A total of 1,313 participants responded to the questionnaires and were, thus, included in the cross-sectional, closely related study (13). Of these, 569 participants agreed to enter the longitudinal phase, with a follow-up assessment planned to take place 18 months after the study inclusion. However, out of these 569 participants, 256 already showed symptoms of burnout at the baseline, cross-sectional assessment (as evaluated by the Oldenburg Burnout Inventory, see below), therefore being excluded from the follow-up. Hence, 313 participants without burnout upon study inclusion were effectively admitted into the longitudinal phase. As such, this can be considered a convenience sample.

Data collection was conducted using a structured electronic form created on *Microsoft Forms.* This form was designed to collect participants’ clinical and epidemiological data, including the presence of any previous chronic disease (comorbidities), along with the following clinical instruments:

- **Oldenburg Burnout Inventory (OLBI):** An instrument adapted and validated in Portuguese that assesses burnout through two subscales: “disengagement” (OLBI-D) and “exhaustion” (OLBI-E). Each subscale consists of eight questions, resulting in a total of 16 items (OLBI Total). Responses are recorded on a five-point Likert scale (5 – strongly agree, 4 – agree, 3 – neither agree nor disagree, 2 – disagree, and 1 – strongly disagree). “Disengagement” reflects a sense of detachment from work, accompanied by cynical and negative attitudes and behaviors related to one’s job. “Exhaustion” refers to feelings of physical fatigue, need for rest, and a sense of overload and/or emptiness in relation to work (14). For this study, we followed the methodology of Delgadillo (15), classifying individuals at or above the mean +1 standard deviation of OLBI Total as having elevated scores, thus indicating burnout. The method was applied using values observed in the Brazilian population (14).
- **Brief Resilient Coping Scale (BRCS):** A unidimensional instrument, adapted and validated in Portuguese, that comprises four items designed to assess the ability to cope adaptively with stress (16). Responses were given on a five- point Likert scale: “5 – Almost always,” “4 – Very frequently,” “3 – Often,” “2 – Occasionally,” and “1 – Almost never”. A score below 13 was classified as “low resilience.”
- **Brief Depression Scale (Patient Health Questionnaire-9 /PHQ-9):** This tool, validated in Brazil (17), consists of nine questions evaluating the frequency of depressive symptoms. A total score of ≥ 9 was considered “high”, therefore, indicative of depressive disorder.
- **Degree of Autonomy in Work-Related Decisions:** A numerical scale was used to measure participants’ perceived level of autonomy in making decisions on their work. Responses ranged from 1 to 10, with 1 meaning “I have no autonomy” and 10 meaning “I have complete autonomy.” Scores ≤ 4 were categorized as a perception of low professional autonomy.
- **Adequacy of the Pedagogical Organization of the Residency Program:** Participants rated their perception regarding the adequacy of the pedagogical structure in their residency programs using a visual numerical scale ranging from 0 to 10, where 0 meant “completely inadequate” and 10 meant “completely adequate.” A score ≤ 5 was defined as indicating perception of poor pedagogical organization.
- **External Work Engagement:** Participants were asked to respond YES or NO to whether they exert professional work outside their residency program.
- **Weekly Work Hours:** Individual were asked about their weekly work hours, with the following options to choose: <24 hours/week, 24 to 60 hours/week, 61 to 90 hours/week, 91 to 120 hours/week, and >120 hours/week.

General characteristics of the studied population were described as absolute and relative frequencies for categorical variables and as measures of central tendency and spread for continuous numerical variables. In bivariate (unadjusted) analyses, associations between several baseline predictor candidates and the outcome of developing burnout during follow up were assessed. For categorical variables, Pearson’s chi-squared tests were applied, with effect sizes estimated as relative risks. For numerical continuous variables, independent-samples t tests (with Welch’s correction for non-homogenous variances when appropriate) and paired-samples t tests as appropriate were used, with effect sizes estimated as differences in means. Baseline characteristics (predictors) showing, in the bivariate analyses, significant association (at p < 0.10) with the subsequent development of burnout (outcome) were carried forward to multivariate (adjusted) analyses, where a log-binomial regression model was applied, with final p-values < 0.05 considered significant. Statistical analyses were conducted in IBM SPSS 25.

This study was approved by the local Research Ethics Committee and the National Research Ethics Committee (CEP/CONEP), registered under the number CAAE 33493920.0.0000.5558 (http://plataformabrasil.saude.gov.br). All participants signed electronic consent forms received via email. The STROBE guideline (18) was used, and all requirements were fully met.

An artificial intelligence-based language model (ChatGPT, OpenAI) was employed to assist in the translation from Portuguese to English and to support the scientific writing and editorial refinement of the manuscript. The authors reviewed and approved all output generated by the AI tool (19).

## Results

In total, 313 participants were included of whom 80.1% (n = 250) were biological females, 52.1% (n=160) were physician trainees, 47.9% (n = 153) non-physician HCP trainees, 97.8% (n= 306) from public healthcare institutions and 90.1% (n = 282) from university hospitals. The mean (SD) age was 28.2 (4.6) years. The general characteristics of the sample are shown in table 1.

**Table 1.**
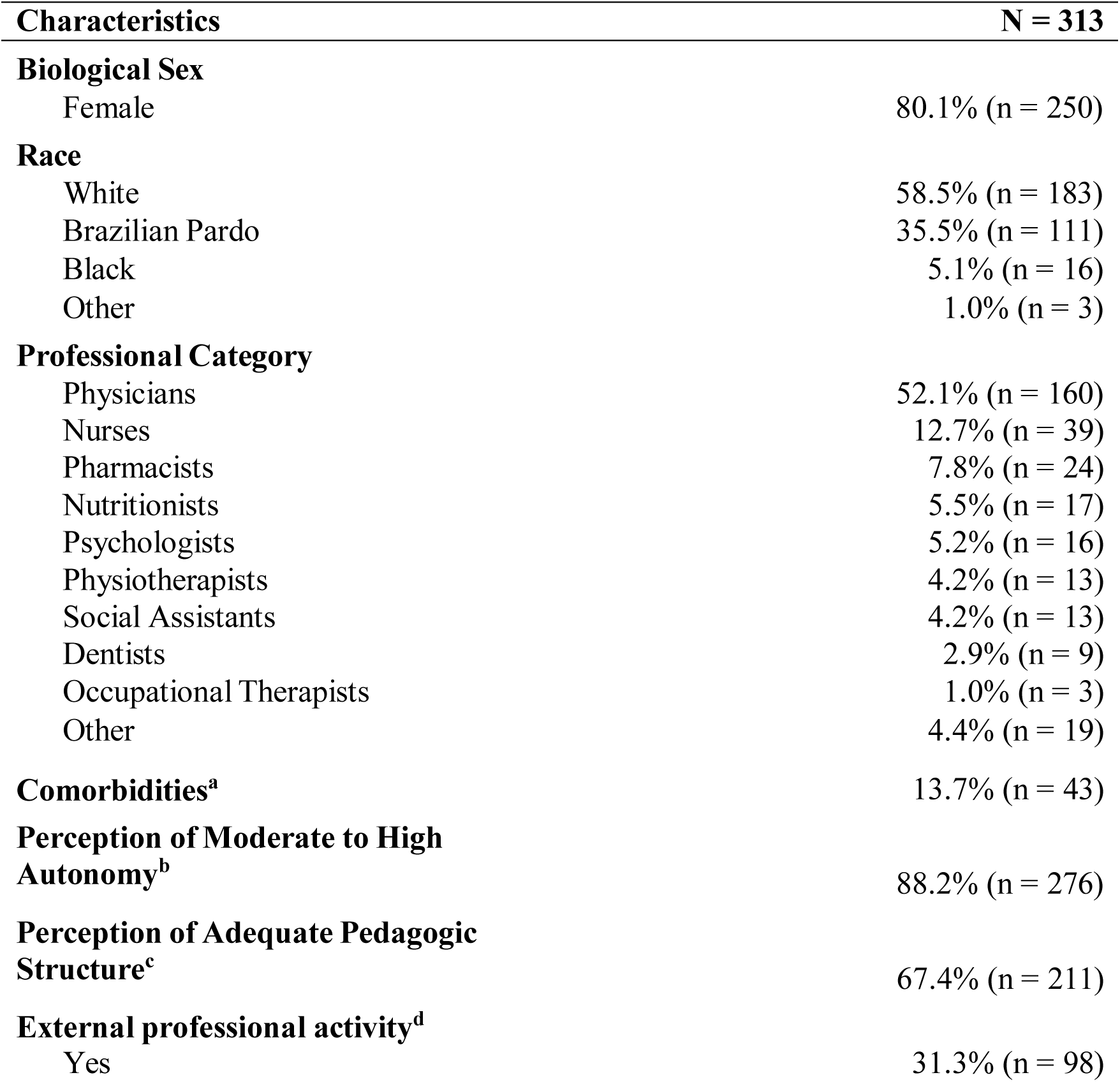
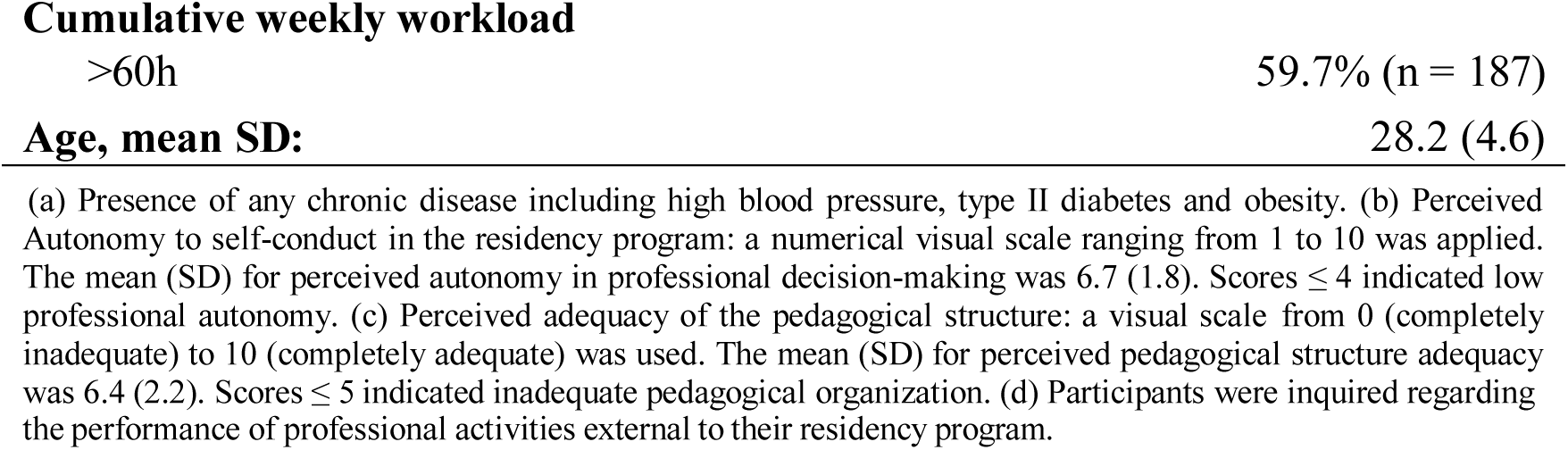
General Characteristics of 313 Healthcare Postgraduate Trainees Working Under a Widespread, High-Demand Sanitary Crisis.

Mean (SD) score on PHQ-9 depression scale was 9.8 (5.3) with 47.9% (n=150) of the participants exhibiting scores ≥ 9, therefore indicating probable depressive disorder. Concerning the BRCS coping resilience scale, the mean SD score was 13.2 (3.5), with 54.3% (n=170) of the participants showing scores < 13, thus classified as having low resilience.

Regarding the OLBI total score, among the 313 participants, the mean (SD) at baseline was 2.8 (0.5). After 18 months of follow-up, the mean (SD) total OLBI score increased to 3.1 (0.7), whereas 30.4% (n = 95) of the participants came to developed symptoms indicative of burnout. Table 2 shows the paired analysis from baseline and follow-up on the OLBI scale.

**Table 2.** Longitudinal changes of burnout scores among 313 healthcare postgraduate trainees during 18 months of follow up, under a high-demand sanitary crisis.

| **OLBI Burnout Scores<sup>a</sup>** | **Baseline** | **Follow-up** | **Differences in means**<br>**[IC 95%]** | **p-value<sup>b</sup>** |
| --- | --- | --- | --- | --- |
| **Disengagement Score,**<br>**mean SD** | 2.4 (0.5) | 2.8 (0.8) | -0.4 [-0.5; -0.4] | <0.001 |
| **Exhaustion Score,**<br>**mean SD** | 3.3 (0.7) | 3.4 (0.7) | -0.2 [-0.2; -0.1] | <0.001 |
| **Total Score, mean SD** | 2.8 (0.5) | 3.1 (0.7) | -0.3 [-0.4; -0.2] | <0.001 |

(a) OLBI – The Oldenburg Burnout Inventory encompassing two subscales (disengagement and exhaustion) and overall score (total). (b) p-values estimated based on paired-samples t tests.

During the 18-month follow-up, 95 new cases of burnout were identified in this population, resulting in an incidence rate of 16.9 per 1,000 person-months (95% CI [13.5–20.3]). Over a one-year period, the incidence rate found was of 202.9 per 1,000 person-years (95% CI [166.5–239.3]).

In bivariate analysis, categorical and continuous numerical variables were examined to identify potential predictors of burnout by assessing their relationship with the outcome (burnout development) over the 18-month follow-up period.

At the 18-month follow-up, healthcare postgraduate trainees who experience burnout were slightly younger than those who did not. The mean (SD) age of trainees without burnout was 28.6 years (5.1), compared to 27.5 years (3.4) among those who exhibited the condition. The onset of burnout was significantly associated with the perception of lower autonomy in decision-making within the training program, perception of an inadequate pedagogical structure at the residency program, as well as the presence of depressive symptoms at baseline. In contrast, variables such as race, sex, comorbidities, type of training program, weekly working hours, professional activity outside the training program, and resilience levels were not significant predictors of burnout. In the analysis of continuous variables, older age (p = 0.029), greater perceived autonomy (p = 0.018), and adequacy of professional training (p = 0.003) were identified as protective factors against burnout (Table 3).

**Table 3.** Associations Between Sociodemographic, Occupational and Mental Health Status and Burnout According to Oldenburg Burnout Inventory (OLBI) Among Healthcare Postgraduate Trainees During a High-Demand Sanitary Crisis: Bivariate Analyses.

| Variable | Burnout<br>(No) | Burnout<br>(Yes) | Association<br>measure<sup>a</sup><br>[IC 95%] | p-value<sup>g</sup> |
| --- | --- | --- | --- | --- |
| **Biological Sex (n = 313)** |  |  |  |  |
| Female, % (n) | 68.8% (172) | 31.2% (78) | 1.14 | 0.563 |
| Male, % (n) | 72.6% (45) | 27.4% (17) | [0.73; 1.78] |  |
| **Race (n = 313)** |  |  |  |  |
| White, % (n) | 67.8% (124) | 32.2% (59) | 1.16 | 0.388 |
| Non-white, % (n) | 72.3% (94) | 27.7% (36) | [0.82;1.65] |  |
| **Comorbidities (n = 313)** |  |  |  |  |
| Yes, % (n) | 58.1% (25) | 41.9% (18) | 1.47 | 0.077 |
| No, % (n) | 71.5% (193) | 28.5% (77) | [0.98; 2.19] |  |
| **Type of Residency Program (n = 313)** |  |  |  |  |
| Non-Medical, % (n) | 67.3% (103) | 32.7% (50) | 1.16 | 0.381 |
| Medical, % (n) | 71.9% (115) | 28.1% (45) | [0.83;1.63] |  |
| **Perceived Autonomy in Self-conduct at work<sup>b</sup> (n = 313)** |  |  |  |  |
| Low, % (n) | 51.4% (19) | 48.6% (18) | 1.74 | 0.010 |
| Moderate/High, % (n) | 72.1% (199) | 27.9% (77) | [1.19;2.55] |  |
| **Perceived Adequacy of Pedagogic Structure<sup>c</sup> (n = 313)** |  |  |  |  |
| Inadequate, % (n) | 53.9% (55) | 46.1% (47) | 2.03 | <0.001 |
| Adequate, % (n) | 77.3% (163) | 22.7% (48) | [1.46;2.80] |  |
| **Cumulative Weekly Workload (n = 313)** |  |  |  |  |
| >60h, % (n) | 68.4% (128) | 31.6% (59) | 1.10 | 0.574 |
| <=60h, % (n) | 71.4% (90) | 28.6% (36) | [0.78;1.56] |  |
| **External professional activity<sup>d</sup> (n = 313)** |  |  |  |  |
| No, % (n) | 67.4% (145) | 32.6% (70) | 1.28 | 0.209 |
| Yes, % (n) | 74.5% (73) | 25.5% (25) | [0.87;1.88] |  |
| **Resilience – BRCS<sup>e</sup> (n = 313)** |  |  |  |  |
| Low, %(n) | 65.3% (111) | 34.7% (59) | 1.38 | 0.068 |
| Moderate/High, % (n) | 74.8% (107) | 25.2% (36) | [0.97;1.96] |  |
| **Depression Score - PHQ9<sup>f</sup> (n = 313)** |  |  |  |  |
| Abnormal, % (n) | 58.0% (87) | 42.0% (63) | 2.14 | <0.001 |
| Normal, % (n) | 80.4% (131) | 19.6% (32) | [1.49;3.08] |  |
| **Age (baseline), mean (SD)** |  |  |  |  |
|  | 28.6 (5.1) | 27.5 (3.4) | -1.1<br>[-2.1; -0.1] | 0.029 |
| **Perceived Autonomy (VAS 1-10), mean (SD)** |  |  |  |  |
|  | 6.9 (1.7) | 6.3 (2.0) | -0.6<br>[-1.0; -0.0] | 0.018 |
| **Perceived Pedagogic structure adequacy (VAS 1-10), mean (SD)** |  |  |  |  |
|  | 6.7 (2.1) | 5.8 (2.3) | -0.9<br>[-1.4; -0.3] | 0.003 |
| **BRCS Resilience score (baseline), mean (SD)** |  |  |  |  |
|  | 13.6 (3.4) | 12.2 (3.6) | -1.4<br>[-2.2; -0.5] | 0.001 |
| **PHQ-9 Depression score**<br>**(baseline), mean (SD)** | 8.8 (5.0) | 12.2 (5.3) | 3.4<br>[2.2; 4.6] | <0.001 |

(a) Association measures for categorical variables, Relative Risks (RR); for continuous numerical variables, differences in means (DM). (b) Perceived Autonomy in self-conduct within the residency program: a numerical visual scale ranging from 1 to 10 was applied. Scores $\leq 4$ indicated low professional autonomy. (c) Perceived adequacy of the pedagogical structure: a visual scale from 0 (completely inadequate) to 10 (completely adequate) was used. Scores $\leq 5$ indicated inadequate pedagogical organization. (d) Participants were inquired regarding the performance of professional activities external to their residency program. (e) BRCS: Brief Resilient Coping Scale. (f) PHQ-9: Patient Health Questionnaire – 9 depression scale. (g) p-values estimated based on Pearson's chi-squared tests for categorical variables and independent-samples t tests (with Welch's correction for non-homogenous variances when appropriate) for numerical continuous variables.

After bivariate analysis, all variables significantly associated with the incidence of burnout (p < 0.05), and two variables with p < 0.10 were included in the multivariate analysis using a log-binomial regression model. This approach aimed to identify which predictors remained significant in increasing the risk of burnout.

In multivariate analysis, the only significant independent predictors of burnout were adequate professional training, which demonstrated a protective effect of approximately 38.6%, and the presence of depressive symptoms at baseline, which increased the risk of burnout by 84.4%. The data from these analyses are presented in Table 4 and in Figure 1.

**Figure 1.**
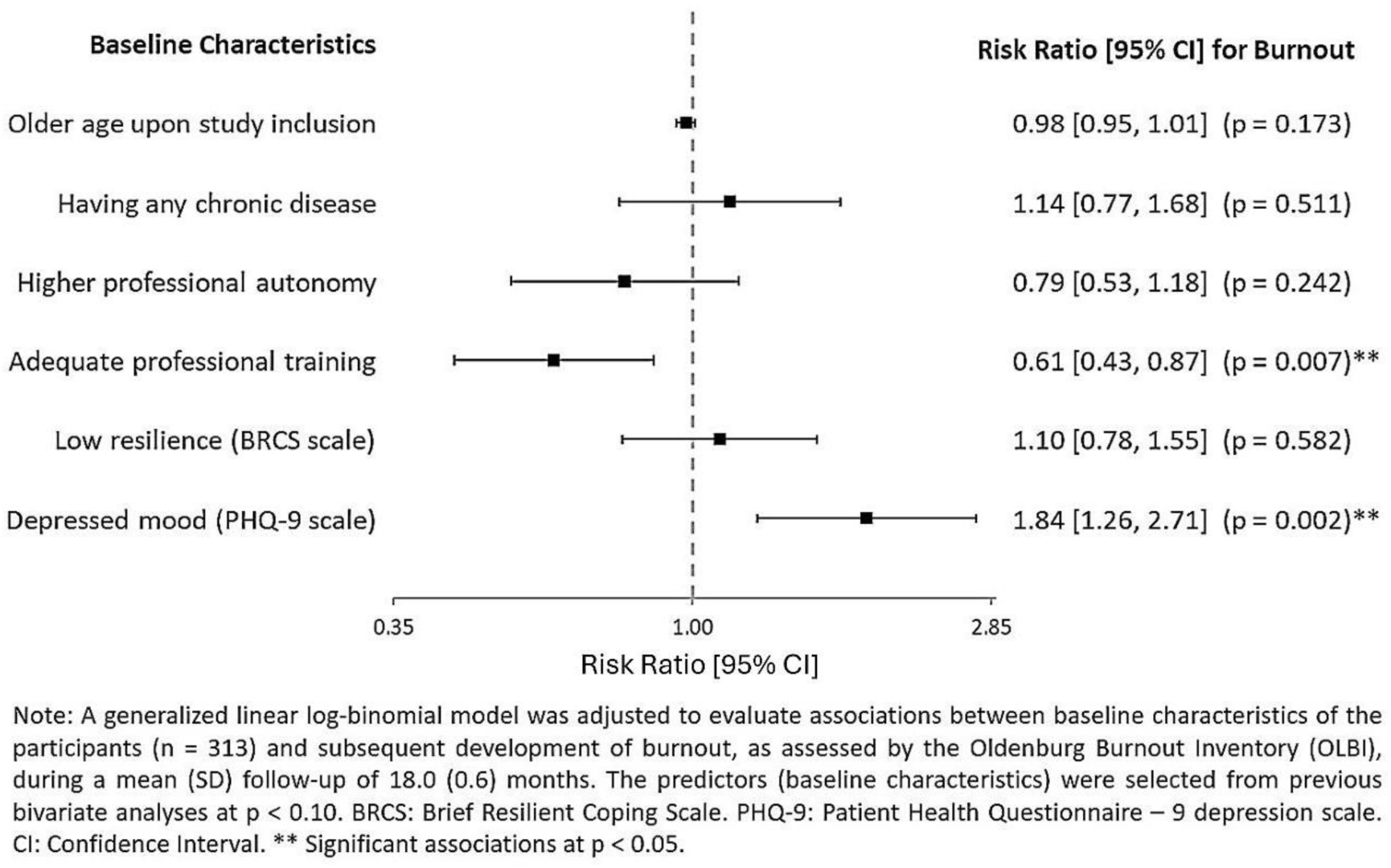
Multivariate analysis of the effect of baseline characteristics to predict burnout development after 18 moth follow-up.

**Table 4.** Multivariate Analysis of the Effect of Baseline Characteristics on the Prediction of Burnout Development After 18 Months of Follow-Up Among Healthcare Trainees During a High-Demand Sanitary Crisis.

| Baseline Characteristics | Risk Ratios (RR) | 95% CI for RR |  | p - value<sup>f</sup> |
| --- | --- | --- | --- | --- |
|  |  | Lower | Upper |  |
| **Older age at study inclusion** | 0.98 | 0.95 | 1.01 | 0.173 |
| **Presence of chronic disease<sup>a</sup>** | 1.14 | 0.77 | 1.68 | 0.511 |
| **Perception of greater professional autonomy<sup>b</sup>** | 0.79 | 0.53 | 1.18 | 0.242 |
| **Perception of better adequacy of pedagogical structure<sup>c</sup>** | 0.61 | 0.43 | 0.87 | 0.007 |
| **Low resilience (BRCS<sup>d</sup> scale – baseline)** | 1.10 | 0.78 | 1.55 | 0.582 |
| **Depressive mood (PHQ-9<sup>e</sup> scale – baseline)** | 1.85 | 1.26 | 2.71 | 0.002 |

(a) Presence of any chronic disease including high blood pressure, type II diabetes and obesity. (b) Perceived Autonomy to self-conduct in the residency program: a numerical visual scale ranging from 1 to 10 was applied. Scores $\leq 4$ indicated low professional autonomy. (c) Perceived adequacy of the pedagogical structure: a visual scale from 0 (completely inadequate) to 10 (completely adequate) was used. Scores $\leq 5$ indicated inadequate pedagogical organization. (d) BRCS: Brief Resilient Coping Scale. (e) PHQ-9: Patient Health Questionnaire – 9 depression scale. (f) A log-binomial regression model was adjusted to estimate the risk ratios and correspondent p-values for all predictors.

## Discussion

This longitudinal study provides critical insights into the incidence and predictors of burnout among postgraduate healthcare trainees—both medical and non-medical—during a widespread, high-demand sanitary crisis. Our results demonstrated the dynamics of individual characteristics and contextual factors in the development of burnout syndrome, thus adding to the global understanding of healthcare professionals well-being under stress.

The study comprised a diverse sample of 313 healthcare postgraduate trainees, predominantly female and mostly young adults. A high proportion of participants reported moderate to high autonomy in their daily professional activities (88.2%) and adequate perception of pedagogic structure in their training programs (67.4%). Nearly 60% reported a cumulative weekly workload exceeding 60 hours. Some of these findings are consistent with those of Amaral et al. (2024) who identified a similarly high prevalence of young (≤28 years, 58.3%) and female (86.1%) residents in their assessment of burnout prevalence among obstetrics and gynecology residents (20)

A high prevalence of depression symptoms (47.9%) and low resilience (54.3%) were identified among these healthcare postgraduate trainees at baseline. A Brazilian cross-sectional study found even higher prevalence of depressive symptoms (61.5%, on PHQ- 9), although with a lower prevalence of low resilience (29%, on BRCS) among medical residents (21). Al-Houqani et al. (2020) reported lower (though still substantial) rates of depressive symptoms (28,8%) among medical residents in Oman (22). Sigal et.al. (2020) reported a significant proportion (33%) of cardiology residents with low resilience levels (23)

Our study revealed a concerning progression of burnout scores among healthcare postgraduate trainees during the follow-up, where 30.4% of the participants came to develop symptoms indicative of burnout. We are not aware of any other study that has evaluated longitudinal changes in burnout scores during the high-demand sanitary crisis posed by the COVID pandemic. Han et al. (2023) in a cross-sectional study reported burnout in 63% of the medical residents investigated (24). Marchalik et al. (2019) found burnout prevalences varying from 26% to 68.2% among urology residents, in a multicenter study conducted in the USA and four European countries (25).

During the 18 months of follow-up, a high incidence of burnout was identified. The incidence rate was 16.9 cases per 1,000 person-months, corresponding to an annual rate of 202.9 cases per 1,000 person-years, underscoring the substantial professional burden within this population.

These findings are consistent with existing literature highlighting the vulnerability of healthcare postgraduate trainees. A longitudinal study conducted by Loyola Netto and Santos (2023) observed a peak in burnout incidence rates during the second half of the second year of residency (26). Although this study did not take place on a sanitary crisis scenario, this temporal pattern suggests a cumulative effect of professional stressors, progressively escalating the risk of burnout over the residency period.

Although longitudinal assessments remain limited, particularly for healthcare postgraduate trainees, the literature consistently documents rising burnout prevalence among healthcare professionals. In Brazilian intensive care unit teams, including physicians and other healthcare providers, burnout prevalence reached up to 64.5% during the second wave of COVID-19, with severe emotional exhaustion observed in 50.5% of individuals (27). In Mexico, a cross-sectional assessment conducted from March 2020 through July 2021 showed that severe burnout among physicians and nurses increased from 30.4% in the pre-pandemic period to 63.2% at the pandemic’s peak (28). Similarly, a pilot study in a Portuguese ICU reported high burnout in 41.3% of nurses, characterized predominantly by marked emotional exhaustion and reduced personal accomplishment (29)

In unadjusted analysis, we observed an association between the occurrence of burnout and younger age. Likewise, in the study by Adam et al. (2018) younger age emerged as the strongest predictor of emotional exhaustion and burnout in Hungarian physician residents (Adam et al., 2018).

Our results showed that perceived low autonomy was identified as a burnout risk factor, with 48.6% of the participants that developed burnout reporting low professional autonomy. This is also demonstrated by Matsuo et al. (2021) on a nationwide online survey of 604 resident doctors in Japan where low autonomy was explicitly identified as a significant stressor contributing to burnout (31). Similarly, Rojo Romeo et al. (2022) on a cross-sectional study, involving 633 medical and surgical residents in France, found that residents reporting lack of autonomy were more than twice likely to experience severe emotional exhaustion (32)

In our findings, low resilience was associated with higher incidence of burnout, with 34.7% of low-resilience participants coming to develop burnout during follow-up. In literature, resilience is repeatedly underscored as a key factor against burnout. Ju et al. (2022) identified self-efficacy (a key component of resilience) as the most significant predictor of lower burnout (33). Similarly, Ho et al. (2021) investigated orthopedic surgery residents in Singapore and found that those experiencing severe burnout exhibited significantly lower resilience scores (34). Consistent with these findings, McCain et al. (2017) reported that, among physicians providing direct patient care, low resilience was associated with the development of burnout (35). Likewise, during the COVID-19 pandemic, a multinational study of the cancer care workforce observed that higher rates of burnout were associated with lower resilience (Cloconi et al., 2023).

In our study, biological sex, race, presence of comorbidities, type of training program, and weekly working hours did not emerge as significant predictors of burnout. Regarding biological sex and weekly working hours, these findings stand in contrast with some prior reports.

Kumar et al. (2016) highlighted biological female sex and workload as key contributors to physician burnout (37). In the same alignment, Adam et al. (2018) found that female residents were more likely to report depersonalization (a key component of burnout), and that greater workload was correlated with depersonalization among female general practitioners(30). Nimer et al. (2021) also reported a significant association between longer working hours and higher burnout levels among Jordan’s residents (38). The lack of association between biological sex, working hours and burnout in our study might be attributable to the long 18-month follow up, under the universally heightened professional distress brought about by the pandemic, potentially reducing the relative importance of these other factors on the occurrence of burnout.

Our findings indicated no association between burnout and the type of healthcare training program. Despite literature review, as of now, we found no studies comparing medical and non-medical healthcare postgraduate trainees regarding the incidence of burnout. However, differences in burnout occurrence have been documented in other studies, when comparing medical and nursing professionals (not trainees). Wozniak et al. (2025) reported, based on a cross-sectional survey, that nurses exhibited higher burnout scores compared to physicians (39). Conversely, Sharma et al. (2008), in their assessment of burnout among colorectal surgeons and clinical nurse specialists, observed that surgeons demonstrated significantly higher levels of depersonalization when compared to their nursing counterparts (Sharma et al., 2008).

The presence of comorbidities did not emerge as a significant predictor of burnout in our study. This contrasts with the existing literature which demonstrates an association between the presence of comorbidities and the occurrence of burnout among healthcare professionals. A study conducted among trainees of Saudi Arabia postgraduate healthcare professions programs observed that healthcare trainees with chronic diseases and those classified as overweight or obese demonstrated higher odds of reporting burnout symptoms (41). Similarly, a study in Taiwan indicated a marginal association between being overweight or obese and a higher risk of elevated personal burnout levels among healthcare workers (42). Furthermore, a multicenter, cross-sectional study conducted in Bangladeshi highlighted that physicians who were either obese or underweight, based on their BMI, faced a heightened risk of experiencing burnout compared to those within the normal weight range (43)

Therefore, the literature consistently shows an association between obesity and burnout. Our study, however, used a broader approach to comorbidity assessment. Because the online design did not allow objective anthropometric measures, obesity could not be analysed separately; all self-reported comorbidities were grouped into a single composite variable. This methodological choice likely accounts for the divergence from previous reports.

In our adjusted analyses, baseline depressive mood emerged as the strongest independent predictor of burnout development, conferring an increase in risk of 84.4%. This finding aligns with prior literature. Chen et al. (2021) identified depression as a major precursor of burnout among healthcare professionals (44). Similarly, Alkhamees et al. (2021) documented a strong association between burnout and depressive symptoms, noting that psychiatry residents experiencing burnout were 8.8 times more likely to exhibit depressive symptoms(45)

Likewise, the perceived provision of adequate professional training emerged as a significant independent predictor, showing a protective effect with an approximately 38.6% reduction in burnout risk. Zhang et al. (2017) found that a mentorship program significantly decreased stress and burnout while improving quality of life among otolaryngology and neck surgery trainees (46). Johnstone et al. (2022) reported that structured mentorship can substantially mitigate burnout among medical urology trainees (47). Together, these findings highlight the importance of educational structures and supportive initiatives in protecting HCP trainees from burnout.

Gregory et al. (2018) evaluated levels of emotional exhaustion (EE) – a core component of burnout – before and after implementing workplace interventions focused on the organization, altering daily practices and protocols to enhance physician’s well-being, finding significant reductions in EE sustained up to six months of follow-up (48). West et al. (2016) advocate for the implementation of comprehensive structural and organizational interventions in healthcare environments aimed at mitigating physician burnout (49). Our findings suggest these interventions should also encompass adequate professional training in order to reduce burnout among healthcare postgraduate trainees.

This study has some limitations. The first is the smaller number of participants who consented to undergo the longitudinal follow-up compared to those enrolled in the cross-sectional phase. Nevertheless, as of now, it is the largest study evaluating the incidence of burnout among healthcare postgraduate trainees.

Another limitation concerns the exclusive use of online data collection without in-person clinical evaluation. However, validated instruments were used to assess burnout and its main predictor-candidates – such as depressive symptoms and resilience – which improves accuracy. Moreover, the online approach proved advantageous, as it enabled broader recruitment.

It is also noteworthy that most participants were affiliated with public university hospitals, which in Brazil generally possess more robust academic and infrastructural framework compared to other public, non-university establishments. This might have attenuated the measured burnout incidence. Therefore, the situation could in fact be even more concerning.

Our study’s unique contribution and strength lie in its approach encompassing both medical and non-medical healthcare trainees and its longitudinal design conducted during a global health crisis. While much of the existing literature focuses exclusively on physician trainees, our study broadens this perspective by showing that these problems are relevant to the full spectrum of healthcare postgraduate trainees. This aligns with findings by Guido et al. (2012), who reported a 27% prevalence of burnout among physicians and other HCP trainees (50).

Understanding the complexity of burnout syndrome and its predictors among trainees is crucial for developing comprehensive, system-wide interventions that will benefit not only trainees, but the entire healthcare workforce of which trainees are an essential component. Addressing burnout is therefore imperative for preserving the integrity of healthcare systems thus maintaining high standards of clinical care. Our findings offer valuable insights to inform strategies aimed at preventing or reducing burnout among healthcare postgraduate trainees and improving their working conditions and institutional support.

## Conclusions

The intense demands of the COVID-19 pandemic resulted in a notably high incidence of burnout among healthcare postgraduate trainees. Depressed mood emerged as the strongest predictor of burnout, while adequate professional training was identified as the most important protective measure among all the variables analyzed. To safeguard the healthcare workforce, providing mental health support and adequate professional training to these trainees during future sanitary crises is strongly recommended.

## Data Availability

All data and related metadata underlying the findings reported in this submitted manuscript is already provided as part of the submitted article.

## Acknowledgments

We extend our gratitude to the University Hospital of Brasília, particularly the Superintendency and the Division of Teaching and Research, as well as EBSERH, for their support on this study.

